# Assessing the feasibility and acceptability of a pre-clinic vital signs assessment in primary care: a pilot study

**DOI:** 10.1101/2024.10.20.24315845

**Authors:** John Broughan, Seán McMahon, Steen Gordon, Nandakumar Ravichandran, Donal Bailey, Jennifer Grant, Geoff McCombe, James Sheil, Walter Cullen

**Affiliations:** Clinical Research Centre, School of Medicine, University College Dublin, Ireland; Wavescope Ireland Ltd., Dublin, Ireland; School of Medicine, University College Dublin, Ireland; Department of Clinical Innovation, Centric Health Primary Care, Ireland; Beacon HealthCheck, Beacon Hospital, Dublin, Ireland

**Keywords:** Feasibility Studies, General Practice, Pilot Projects, Primary Health Care, Vital Signs

## Abstract

**Background:** Vital signs assessment can be crucial. However, such assessments are time-consuming and so are not always prioritised. Measuring vital signs before doctor visits may therefore be an effective and efficient strategy. We piloted a pre-clinic vital signs assessment (PCVSA) within a primary care centre to determine its feasibility and acceptability.

**Methods:** A mixed methods cross-sectional design was piloted. Study participants included adult patients and practice staff. Patients had vital signs assessed by a Primary Care Assistant before GP visits. Data collected concerned participants’ study engagement, the timings of PCVSA / GP visits, and surveys / interviews investigating participants’ experiences.

**Results:** Sixteen patients and four staff participated. The mean PCVSA was 2mins23secs (SD = 38.8) and the mean GP visit was 9mins21secs (SD = 252.4). Patients said the PCVSA was a ‘Positive experience’ (87%), ‘Helpful’ (81%), ‘Valuable’ (44%), and ‘Interesting’ (38%). The GP said the PCVSA was either ‘Helpful’ (n=8, 54%) or ‘Extremely Helpful’ (n=7, 47%) in each of their consultations, and that it improved engagement with 80% of patients, allowed them to spend more time gaining understanding of the conditions of 93% of patients, and enhanced productivity in 73% of consultations. The GP strongly agreed that collecting PCVSA data before appointments would benefit patients over time. Qualitative interviews with practice staff yielded three themes: (1) Improved patient engagement and efficient consultation, (2) Time-saving potential, and (3) Practicing in general practice and associated challenges.

**Conclusion:** The PCVSA pilot showed good feasibility and acceptability as indicated by high participant engagement, short PCVSA and GP visit times (albeit GP visit times did not measure non-patient facing clinical activity), and positive feedback from patients and staff. Introducing PCVSA in healthcare settings may have potential in terms of improving the standard and efficiency of care.

**Author summary:** Checking vital signs is important for patient care, as it helps doctors assess, monitor, and make clinical decisions. However, it is often overlooked to save time for other tasks considered more urgent. A new approach called ‘pre-clinic vital signs assessment’ (PCVSA), where patients’ vital signs are measured before their one-on-one visit with the doctor, may help address this issue. This study tested the PCVSA at a primary care centre in Dublin, Ireland, to see if it could improve care quality, save time, and be accepted by both staff and patients. On average, the PCVSA took 2 minutes and 23 seconds, while the typical doctor visit lasted 9 minutes and 21 seconds. Most patients and staff saw the PCVSA as useful, highlighting its benefits for better patient-doctor communication and timesaving. Some concerns were raised about the extra demand it might place on clinic resources. Overall, the findings suggest that PCVSA could positively impact the quality and efficiency of care in primary care settings, with future research needed to explore its benefits in larger and more varied clinics.

## Background

Assessment of patients’ vital signs is an important component of healthcare delivery, particularly among acute patients. Vital signs assessment allows clinicians to investigate and monitor patient health in an objective manner. Patient safety is also an inevitable concern in healthcare and studies have reported that vital signs assessment acts as a tool to improve patient safety in the medical unit reducing the risk of adverse events and provide better care for patients [1-3]. Thus, regular vital signs assessment can be important to ensure efficient and potentially critical clinical follow-up and intervention for patients as necessary [4].

However, despite vital signs assessment’s well-established benefits, because of global healthcare professional shortages, vital signs screening is often impractical in busy modern healthcare settings [5]. Adding vital signs assessments in the manner that they are conducted today to every consultation would be problematic given the well documented existing pressures on staffing and clinical flow [6]. A survey of 25 GPs conducted by Wavescope Ltd. from February 2023 to January 2024 (Supplementary material) found that GPs take patients’ vital signs in around 30% of consultations, and that they could spend approximately three minutes taking vital signs in each of these. Accounting for assumptions regarding hours worked per day (8 hrs) and weeks worked per year (46 weeks), our calculations indicate that full-time GPs in Ireland spend approximately 110 hours per year just taking patients’ vital sign measurements.

Measurement of vital signs can also result in “white coat hypertension”, a phenomenon characterised by elevated blood pressure measurements and other artefacts due to stress experienced by patients in the presence of healthcare professionals [7]. Further, research indicates that vital signs assessments are often undervalued by clinicians, not recorded faithfully or frequently enough, and the findings of vital signs assessments may be disregarded as unimportant [2]. For instance, a retrospective analysis by Hayes et al. of patients, in examinations where international guidelines recommended a vital signs assessment, revealed that 98% of patients had substandard vital signs records [8].

The worst-case scenario consequence of poor adherence to optimal vital signs assessment procedures is a greater probability of medical problems among patients not being detected by clinicians and patient health deteriorating as a result, potentially to a severe degree among patients with acute health problems [9]. To that end, this study intended to address these issues by introducing a Pre-Clinic Vital Signs Assessment (PCVSA) procedure in a busy primary healthcare setting. Each patient’s vitals assessments were conducted by a Primary Care Assistant (PCA) while they waited to see the general practitioner (GP). Their vital signs data were produced in a report sent to the GP before they saw the patient, thus allowing them to maximise the time they spend on clinical decision making. We hypothesised that the PCVSA would be deemed feasible from the perspectives of key stakeholders in the primary care setting, namely the GP, clinical support staff and patients.

## Methods

### Research design

A mixed methods cross-sectional design involving calculation of PCVSA times and stakeholder questionnaire / qualitative interviews was piloted. The PCVSA intervention was assessed in terms of its feasibility and acceptability using Bowen et al.’s feasibility study framework [10]. As per this framework, the PCVSA was evaluated according to its acceptability, its level of demand among stakeholders, practicality, and capacity for implementation, integration, and expansion within busy primary healthcare settings.

### Setting, participants, and sampling

The study was conducted within a large, busy, primary healthcare centre in an urban suburb of Dublin City, Ireland during the 9am to 12.30pm shift each day from July 8th to 10th, 2024. Study participants included key staff working at the practice, as well as a target of 20 adult patients attending the practice for their routine GP visit appointments. Research regarding calculating pilot study sample sizes indicates that this size is suitable for a pilot feasibility study [11]. Inclusion and exclusion criteria for patients participating in the study were formulated and communicated to the practice’s clinical staff before recruitment of patients as participants. These criteria are outlined below:

Inclusion criteria:

- Patients over age 18.
- Patients able to provide written informed consent to participation (or to be supported to provide informed consent as per the Assisted Decision Making (Capacity) Act 2015).
- Patients due to see the doctor with a new or ongoing physical health concern or problem.

Exclusion criteria:

- Patients whom the clinical staff (GP, PCA) determine as too unwell to wait to see a clinician; instead, being “fast tracked” into assessment.

### Recruitment

#### Patient recruitment

Patients participating in the study were recruited as they attend the practice for their pre-scheduled, non-study related, GP visit appointments. On presenting to the clinic for check in, the patients were informed by the practice receptionist that the clinic is running a pilot study which is intended to assess the feasibility and acceptability of introducing a PCVSA. The patients were provided with a study information sheet which included all relevant information on the study and were given the opportunity to read the material. The PCA was there to assist patients with these documents and members of the study team were available onsite to provide more detail and answer questions from potential participants if requested. Patients were also given the opportunity to take home the study information sheet to think about potentially participating in the study and then provide informed consent on their next visit if the study was still in progress. Patients that wished to participate were provided with a consent form to review and sign before taking part in the study.

#### Staff recruitment

Staff selected by the practice management team were invited to participate in the study also. If they wished to take part, staff were provided by the research team with a participant information leaflet explaining the study and were offered the opportunity to discuss the study with the research team. Staff also had to sign a consent form to participate in the study.

### Procedure

Each patient was provided with a unique Radio Frequency Identification (RFID) sticker that was attached to a card [12]. A custom RFID code [13] was designed specifically for the study and this automatically collected RFID data and the time when the RFID sticker was presented to RFID readers [14]. The PCA called each patient for their vital signs assessment and the patients scanned their RFID card at a dedicated vital signs assessment station in the PCA’s private office. The PCA then measured the patient’s vital signs (temp, BP, Oxygen saturation, heart rate and respiration rate) using a vital signs measurement machine (Welch Allyn Connex® Vital Signs Monitor 6000 Series supplied by MDI Ireland ltd). Provisions were made to warrant up to three vital signs assessments at 5, 10 and 15 minutes if results were outside the range of normal. The PCA then placed the RFID card on the RFID scanner to log the time of the examination initiation and termination.

The patients returned to the waiting area, and their vital signs were digitally transmitted to the doctor via practice-based management software (Socrates) for review prior to them being admitted to the doctor’s consultation room. The PCA requested that once in the GP’s consultation room, the patient scan their RFID card before the beginning of their visit to the GP’s office and again when the face-to-face meeting is complete. The GP and patients completed brief questionnaires querying their experience of the PCVSA consultation. The GP also completed an additional “Doctor Satisfaction Questionnaire” at the end of the entire pilot study to gather their overall impressions of the study and any additional information. All questionnaires were developed by Wavescope Ltd. in collaboration with colleagues from Afortiori Development, Ulster University, and UCD. Lastly, qualitative interviews were conducted with practice staff to investigate their experiences of the study in more detail. The study procedure is outlined in Figure 1 below. Study questionnaires and the qualitative interview schedule are included in this paper’s supplementary material.

**Figure 1.**
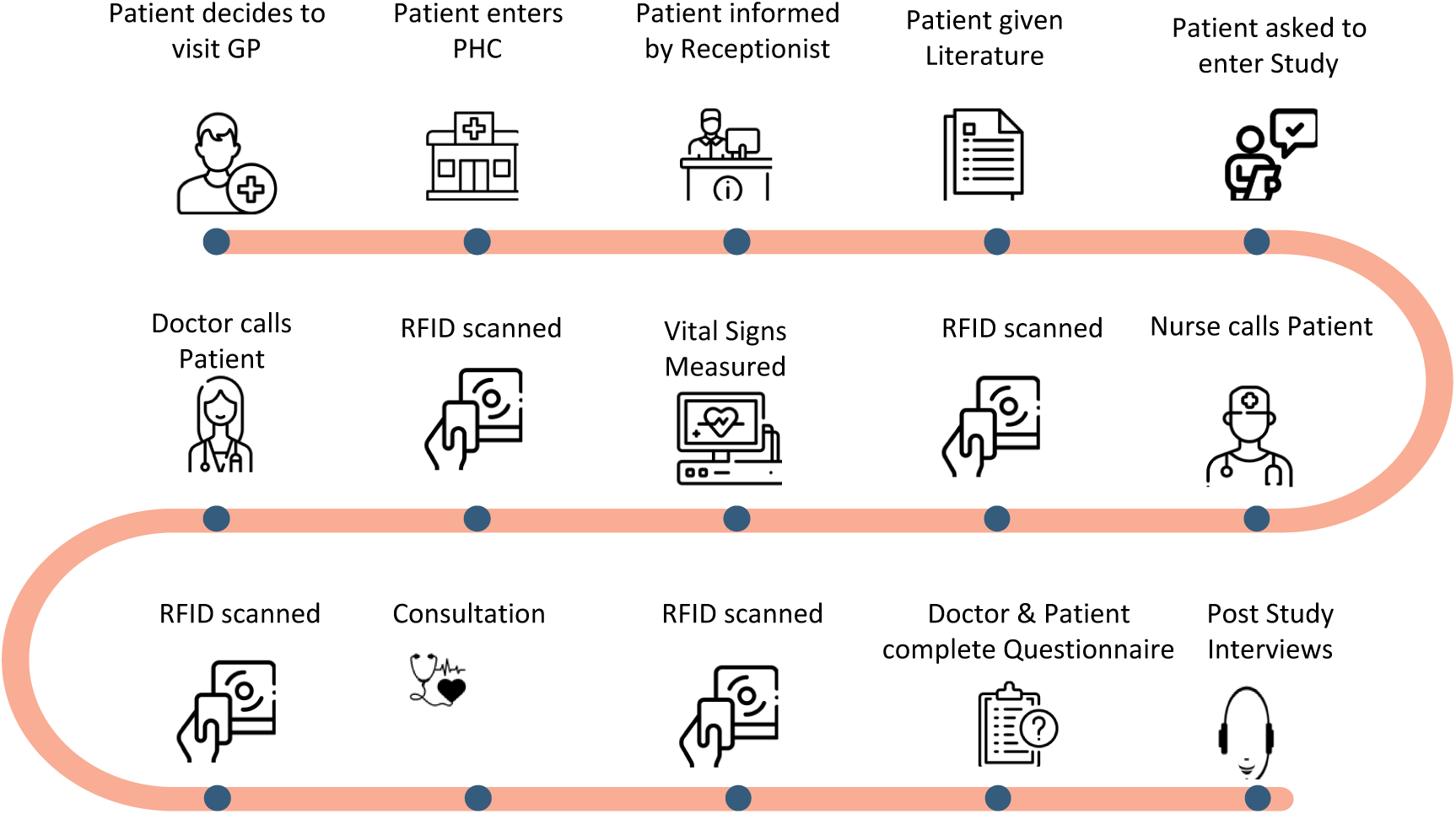
Study procedure.

### Data collection

Study engagement data (no. patients invited to participate, no. that participated, no. that completed the study), were collected to determine participant recruitment and retention rates. The exact timings of each stage of patients’ visits were recorded using the RFID technology with patients scanning their RFID cards before beginning and completing the face-to-face PCVSA and GP appointments. While patients’ vital signs data (temp, BP, Oxygen saturation, heart rate and respiration rate) were recorded, this data was retained by the clinic, was not accessible to the research team, and was not used in the study.

Both patients and the GP completed brief questionnaires following each patient interaction asking them questions about their experience of the PCVSA. These included a ‘Doctor Assessment Questionnaire’ completed by the GP for each patient enrolled in the study and ‘Patient Assessment Questionnaires’ completed by patients themselves. The ‘Patient Assessment Questionnaire’ included open and closed items querying patients’ demographics (age range [18 -40, 40 -65, 65+] & gender), and whether they felt the PCVSA was a positive experience, one that they would be happy to take part in at every GP consultation, and one that was helpful, valuable, or interesting. The ‘Doctor Assessment Questionnaire’ also included open and closed statements asking about how helpful it was having the PCVSA was before the start of their face-to-face interaction with the patients, as well as the extent to which they agreed with statements saying that the PCVSA improved their engagement with patients, allowed them to spend more time understanding the patient’s condition, and offered time or productivity gains during the consultation. At the end of the study, the GP also completed another brief questionnaire (‘Doctor Satisfaction Questionnaire’) asking them to indicate overall, how beneficial they felt the PCVSA was, which collected vital signs they found to be most useful, what other pre-clinic measurements they would like to see taken in future, and if they would accept vital signs data generated by patients personal wearable devices.

Lastly, key practice staff participated in in-depth semi-structured qualitative interviews once their involvement in the study was complete to acquire further insight into their experiences of the study. This data was collected via audio recordings of the interviews which were subsequently transcribed and pseudonymised for analysis. As per Bowen et al.’s feasibility study framework, the qualitative interview topic guide focused on areas pertaining to the study’s acceptability, practicality, adaptability, likely efficacy, the demand for PCVSAs, and the PCVSA’s capacity being implemented in, integrated within, and expanded to other healthcare settings.

### Data analysis

Descriptive statistics (frequency counts, percentages, timings of consultation stages, measures of central tendency and variation) were used to analyse quantitative data including study engagement, age, gender, PCA/GP face-to-face visit timing, and closed response questionnaire data. Qualitative coding techniques were used to analyse open response questionnaire data and analysis of participants’ qualitative interviews was guided by Braun and Clarke’s ‘Reflexive Thematic Analysis’ approach [15].

### Ethics

Ethical approval for the study was obtained from the UCD Human Research Ethics Committee (LS-24-36-Broughan-Cullen). In line with open science practices, the study hypotheses and planned analyses were preregistered on the AsPredicted website [16].

## Results

### Study engagement and participant characteristics

Thirty-six patients were scheduled for appointments with the GP during the study, 12 on each day. Of these, 14 (39%) did not participate for reasons including appointment cancellations, language barriers, and not meeting the eligibility criteria (i.e., being under 18 years of age). Sixteen of the remaining 22 patients (72%) agreed to, consented to, participated in, and completed the study. The sixteen patients comprised of 12 females (75%) and four males (25%), and most patients fell within the 40–64-year age range (56%) (see Table 1). As per prior agreements with site management personnel, four practice staff members (GP, PCA, Receptionist, and Practice Manager) selected by the practice management team were invited to take part in the study and all four staff consented to doing so. The GP in the study was male and the three other staff participants were female.

**Table 1.**
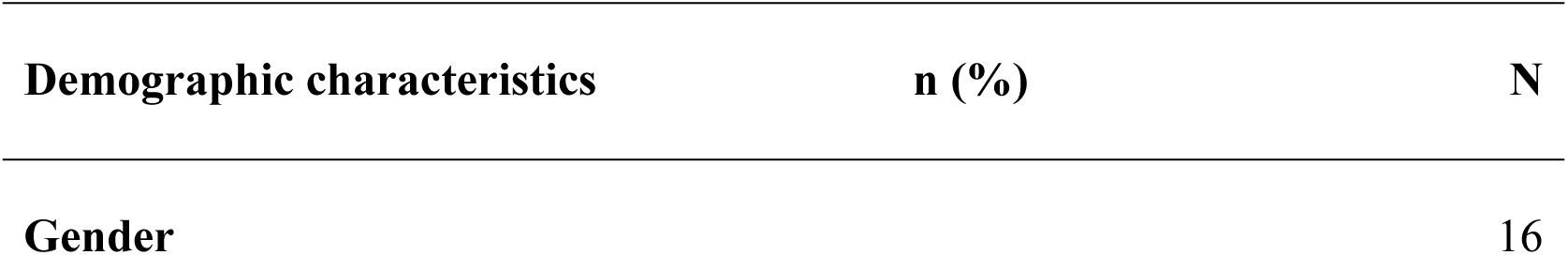
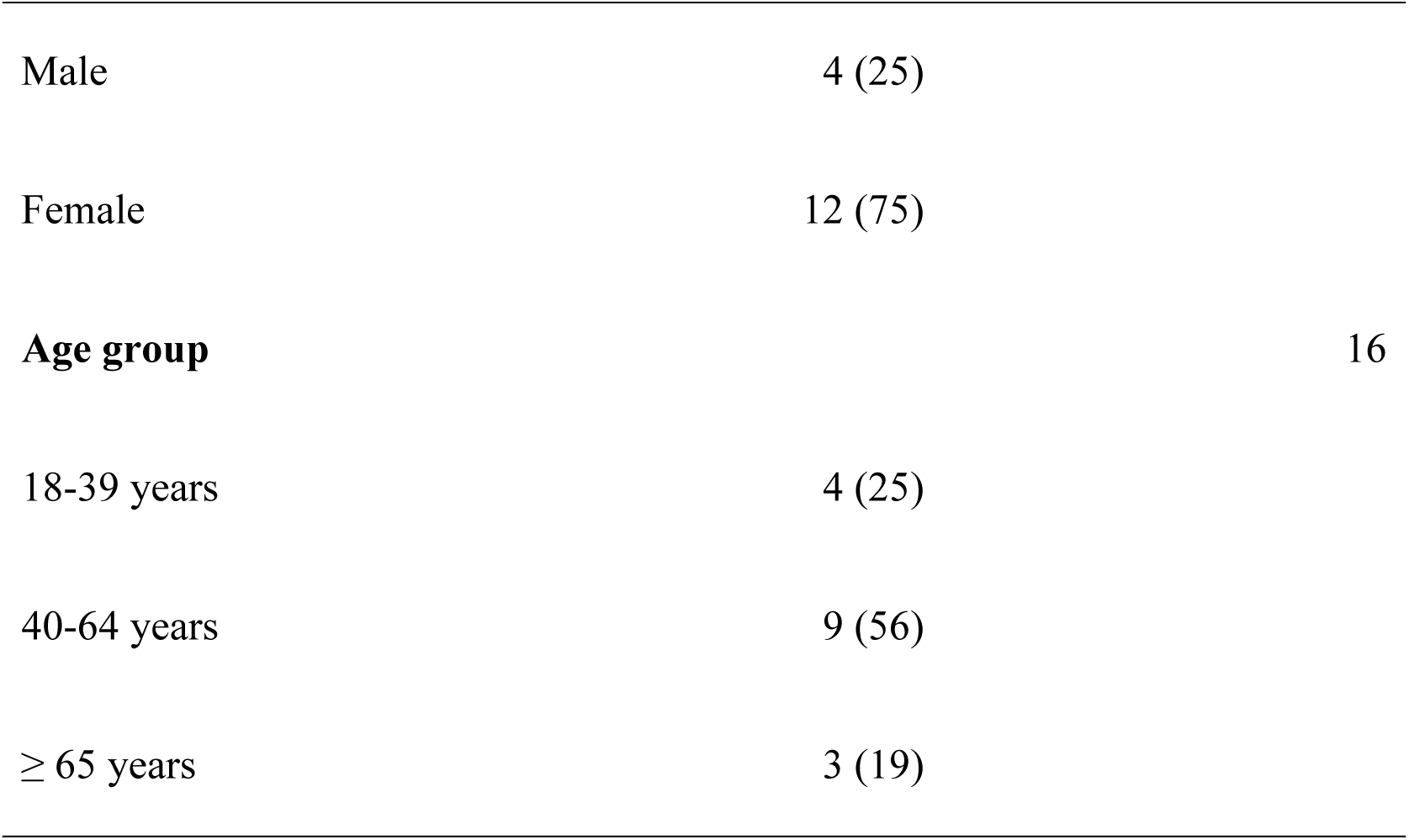
Patients’ demographic characteristics.

### Timings of PCVSA and GP face-to-face visits

The mean duration spent by the PCA on the vital signs assessment (n=16) was reported as 142.6 seconds (2mins 23secs) and the mean average duration of the face-to-face GP visit (n=16) was 560.5 seconds (9mins21secs) (see Table 2).

**Table 2.** Time spent during the PCVSA and GP interactions (n=16).

| **Time spent** | **Mean (SD)** | **Median (P25 – P75)** |
| --- | --- | --- |
| ***PCVSA*** | 2mins 23secs (38.8) | 2mins 36secs (108.2 – 169.8) |
| ***GP visit*** | 9mins 21secs (252.4) | 8mins 59secs (364.3 – 671.7) |

### Responses to the ‘Patient Assessment Questionnaire’

Most patients strongly agreed that it was a positive experience to take vital signs assessments prior to their face-to-face visit with the GP (88%) (see Table 3). All 16 patients said ‘Yes’ to a question asking if they would be happy for their GP to collect data on their vital signs during every consultation. Patients were asked if they found the PCVSA to be ‘Helpful’, ‘Valuable’, or ‘Interesting’. Thirteen of 16 patients (81%) said it was ‘Helpful’, seven of 16 (44%) said ‘Valuable’, and six of 16 (38%) said ‘Interesting’.

**Table 3.** Having my vital signs assessment taken prior to meeting the GP was a positive experience (n=16).

| Characteristics | n (%) |  |  |  |  |
| --- | --- | --- | --- | --- | --- |
|  | Strongly disagree | Disagree | Neutral | Agree | Strongly Agree |
| Having my vital signs assessment taken prior to meeting the GP was a positive experience. | 0 (0) | 0 (0) | 0 (0) | 2 (12) | 14 (88) |

### GP responses to the ‘Doctor Assessment Questionnaire’

Fifteen of 16 possible ‘Doctor Assessment Questionnaire’ responses were collected. The responses indicated that the GP felt having the PCVSA before meeting patients was either ‘Helpful’ (n=8, 53.8%) or ‘Extremely Helpful’ (n=7, 46.7%) (see Table 4).

**Table 4.** What was the value of having the PCVSA information at the before meeting patients? (n=15)

| What was the value of having the PCVSA info before meeting patients? | Unhelpful | No impact at all | Helpful | Extremely helpful |
| --- | --- | --- | --- | --- |
|  | 0(0%) | 0(0%) | 8(53%) | 7(47%) |

The GP strongly agreed that the PCVSA improved engagement with 80% of the patients. They also strongly agreed that PCVSA allowed them to spend more time gaining understanding of the conditions of most of the patients (93%), and strongly agreed that the PCVSA enhanced productivity gain in 73% of the consultations (see Table 5).

**Table 5.** Did having the vital signs assessment prior to meeting the patients… (see below) (n=15)

| Characteristics | n (%) |  |  |  |  |
| --- | --- | --- | --- | --- | --- |
|  | Strongly disagree | Disagree | Neutral | Agree | Strongly agree |
| Improve how you engage with the patient? | 0 (0) | 0 (0) | 0 (0) | 3 (20) | 12 (80) |
| Allow you to spend more time gaining understanding of patient's condition? | 0 (0) | 0 (0) | 0 (0) | 1 (7) | 14 (93) |
| Offer any time or productivity gain in the consultation? | 0 (0) | 0 (0) | 0 (0) | 4 (27) | 11 (73) |

### GP responses to the ‘Doctor Satisfaction Questionnaire’ (n=1)

The GP indicated that the PCVSA was beneficial. On a scale of ‘Strongly disagree’ - ‘Agree’- ‘Neutral’ - ‘Agree’ - ‘Strongly agree’, the GP strongly agreed that collecting PCVSA data in advance of meeting patients would benefit patients over time. When asked which (if any) of five vital signs measurements (temperature, heart rate, respiration, blood pressure, oxygen saturation (SpO2)) they found to be most useful, the GP indicated that all measurements were equally useful. Additionally, the GP suggested incorporating additional measurements into pre-clinic assessments such as height and weight for chronic patients, and ECGs for certain patients. However, the GP was uncertain about accepting vital signs data from patients’ own wearable devices.

### Findings from qualitative interviews with practice staff

In-depth semi-structured qualitative interviews were conducted with practice staff members. These included the GP, PCA, Receptionist and Practice Manager. The following themes were identified from the interviews: (1) *Improved patient engagement* and *efficient consultation*, (2) *Time-saving potential*, and (3) *Practicing in general practice and associated challenges*.

#### Improved quality of doctor / patient engagement

All participants agreed that the PCVSA initiative improved patient engagement, allowing more time to understand the patient’s condition and enhancing the productivity of consultations. The GP highlighted that while patient engagement and consultation productivity varied from case to case, the initiative generally allowed more attention to patients, thereby improving GP-patient engagement.

The GP commented on patient engagement:

> *“It’s difficult to say (whether the PCVSA improved patient engagement), it differs case to case but yeah, you get more time and more attention.”*

The practice of having both the GP and the PCA check the patient’s vital signs before they met the GP was seen as providing extra care, with the PCA being particularly perceived as a sympathetic and empathetic support for patients, leading to improved consultations. The staff also highlighted that in their view, the PCVSA helped patients feel more relaxed, reducing potential nervousness and anxiety (i.e., ‘White Coat Syndrome’) that could lead to improper vital signs measurement. Participants reported that patients were happy with having their vitals taken before they met the GP.

The practice manager commented:

> *“…they (patients) feel they get the extra care, because there’s two health professionals looking after them as well…”*

The participants also mentioned that knowing vital signs beforehand could hypothetically allow staff to alert the GP to any alarming conditions.

The PCA commented:

> *“…it was great for us and for the patient to know that their vital signs were all fine or if there was any problems before they had even gone into the GP, where he knows if there was anything flagging up, if they had a fast heart rate or their BP was high, that they know that he could see that before he even called the patient in, so we kind of had an idea of what was coming in to him…*

#### Time saving potential

Having vital signs taken before the face-to-face meeting with the GP meant that the GP had all necessary information readily available, saving time as they could immediately proceed with patient facing consultation duties. Participants highlighted that the study process was quick and easy, potentially allowing for PCVSA to be implemented across two to three GPs working in parallel on a single day. This efficiency enabled them to meet with more than 16 patients per day easily, despite some language barriers in consenting for the study.

The PCA commented:

> *“I would a million percent jump on board with the PCVSA study. I could have definitely managed two GPs in one day. We saw 16 patients, but I would have liked to see more. Unfortunately, some patients were children, some faced language barriers, and some needed to take information home to translate. Additionally, some elderly patients could only handle limited participation. However, many patients, particularly those in their mid-20s to 40s, were very interested. With their enthusiasm, I think I could have managed two or three GPs seeing patients simultaneously.”*

While time savings varied by case, there was a consensus that the initiative saved a considerable amount of time.

The GP commented:

> *“Each case is different. Some cases take more time, some cases less time, you know… but as a general rule, if you ask me, yeah, the vitals will save time.”*

#### Practicing in General Practice and Associated challenges

While some participants noted that similar initiatives had not been undertaken before, others mentioned implementing comparable measures during COVID-19 as an infection prevention and control measure. These measures were typically applied to acute patients rather than in general practice appointments. All participants agreed on the necessity of initiatives like PCVSA in primary care and general practice, indicating they would recommend it to colleagues.

The PCA commented:

> *“I do think it’s important and I do think that if it did come into the practices, it would really help a lot and it would catch a lot of them, of high blood pressures or fast heart rates or, you know, and then that you’re able to get them under control as well and a GP to obviously, you know, starting it with a medication if needed or looking for the testing if needed or just to keep an eye every so many weeks or so many days and I think it was a really, really good idea, having a practice, especially before going into a GP.”*

One participant emphasised that in the long term, 30-40% of people might not get their vitals checked as part of routine care, making it crucial to implement this initiative to avoid future problems. The GP also highlighted that the PCVSA would be particularly beneficial for patients with acute conditions, (e.g., exacerbation of COPD, respiratory tract infections, fever of unknown origin) although it may not be as useful for those with general ailments. Implementing the initiative in general practice was considered beneficial overall.

The GP commented:

> *“You know, mostly it is, but in some instances, as I mentioned, you know, if you have a trauma or anything like that, you know, the vitals are not as important as in acute cases. Just in acute cases. I mean, it saves time, you know, in acute cases, you know, so you can be more productive. Yeah. And even if it’s not acute sort of a thing, I mean, there’s no harm in doing it, if you have something in front of you.”*

One of the challenges highlighted by participants was the need for additional staff to perform PCVSA alongside other nursing duties.

The GP commented:

> *“I mean, generally, the PCVSA will help, you know, but then you need the manpower and extra staff to do the vital signs, besides their own things, especially the nurse (PCA).”*

The study was found to be interesting, with both patients and staff expressing satisfaction in participating. Some challenges were noted, particularly related to language barriers during consenting process.

The receptionist commented:

> *“Most people on board were happy, but there were some people that we found issues with, like, language was a barrier. So, I didn’t have time to sit down and explain the study fully to them…So, they didn’t want to sign-up for the study because they didn’t understand (what it involved). Yeah. So, it’s hard to explain. (We) needed a little bit more time explaining them about the study.”*

## Discussion

### Summary of key findings

This study aimed to determine the feasibility of piloting a PCVSA initiative in a busy primary healthcare setting in Dublin, Ireland. Good feasibility was observed as evidenced by the short time durations spent by patients during their PCA assessments and subsequent face-to-face meetings with GPs, as well as high levels of acceptability for the PCVSA among participating patients and practice staff completing post-PCVSA questionnaires and qualitative interviews. While 14 (39%) of patients scheduled for appointments were not eligible to participate because they cancelled GP appointments, did not meet the predefined eligibility criteria, or were deemed by practice staff to not have necessary English language skills to sufficiently comprehend the study’s participation information sheet or consent form, there were good levels of study participation and completion among eligible patients attending the practice and allocated staff.

As for the study’s acceptability, frequently observed positive impacts of the PCVSA included a desire among patients and staff to see PCVSA being implemented within standard care practices, perceived PCVSA related improvements in the quality and efficiency of care provided, and perceived enhancements regarding staff / patient relationships and communication. The most notable feasibility issues that were identified included the additional workload that routine PCVSA could have on practice staff, and the necessity for strategies in future studies to overcome language barriers impeding participation in the study for some patients.

### Comparing with existing literature

Research indicates that vital signs assessment can be important to ensure effective clinical evaluation, providing crucial information about a patient’s health status and playing a key role in triage, prevention, and patient safety within healthcare settings, albeit particularly for vulnerable patients and patients requiring emergency and hospital care where the bulk of research has been conducted to date [17-21]. Our study points to the importance of PCVSA in a general practice setting, with the findings of this study emphasising the perceived value among practice staff of PCVSA in saving time and improving patient engagement and consultation quality.

The average time reported by the PCA for the conducting of PCVSA in our study (2.4 minutes) is notably shorter than the 3.75 minutes reported by a similar study [95% CI 3.53 to 3.9] [22]. However, the study mentioned was conducted in a hospital rather than primary care setting, and systematic review research mentions that meaningfully comparing timings for vital signs assessments between studies is difficult or even impossible because of variations in the methods of vital signs assessment between studies [3]. It is also important to note that the average GP visit time reported in this study (9mins21secs) only captures the GP’s patient facing clinical work and should not be confused with complete consultation time which previous research by Crosbie et al. (2020) and Pierse et al. (2019) calculated to range from 14.1mins to 14mins36secs [23, 24].

Our study highlights factors such as White Coat Syndrome, patient nervousness, and anxiety as potential contributors to inaccuracies in vital signs measurements. This aligns with existing research that identifies these factors as common sources of inaccuracy in vital signs assessment [25-27]. The findings show that the GP in this study ranked temperature, heart rate, respiration rate, blood pressure, and oxygen saturation as the most useful vital signs. This aligns with the literature, albeit not based within primary care settings, that emphasises these parameters as critical indicators of patient health [28-32]. Additionally, our study supports recommendations from systematic reviews that advocate for the inclusion of pulse oximetry in vital signs measurements to improve patient care and outcomes [33].

The study by Ullah et al. (2022) reported on the increased workload associated with vital signs measurement in general practice, a very important consideration. However, our study’s findings demonstrate that the use of PCVSA, despite concerns about support staff workload and language barriers during the consenting process, did not hinder the PCA in their efforts to manage 16 patients and support other GPs working in the practice that day simultaneously.

This led to a perceived reduction in times spent during face-to-face GP visit times and GP workload among participating staff, as well as improved patient experiences and satisfaction. Elliott et al. (2024) identified gaps in the rigorous assessment of vital signs, emphasising the need for consistent and thorough measurement to ensure patient safety and effective surveillance in acute settings [34]. Our study’s findings provide support for Elliot et al.’s position in a primary care context. The positive feedback from both patients and staff underscored the value of PCVSA in improving patient engagement, consultation efficiency, and overall satisfaction in this setting. Having said this, there remains a need for further testing of PCVSA in general practice on a larger scale across diverse contexts to fully determine the value of Elliot et al.’s stance.

#### Implications for research, policy and practice

This study’s findings indicate that PCVSA could have several positive implications for clinical practice. The findings suggest that in general practice settings, PCVSA may lead to improvements regarding the quality of patient care, identification of health problems, operational workflow and time management, patient and staff satisfaction, and relationships and communication between patients and clinical staff. It is also possible that the benefits of PCVSA may extend to other care settings such as nursing homes or hospitals, albeit evaluation of PCVSA studies in these settings is required before such claims can be made with confidence. A barrier to routinely conducting vital signs assessments is the extra time and workload that assessments can bring [5]. A PCVSA that does not require direct clinician, or even healthcare staff involvement (e.g., PCVSA from clinically validated personal wearable devices), may overcome this impediment, thus increasing the likelihood that busy clinical staff are more likely to comply with the vital signs assessment recommendations advised by practice and policy influencers and decision makers. From a research perspective, the main implication of this study’s findings is an evidenced need for continued research examining the potential of PCVSA, in primary care as well as in other care contexts. Larger-scale feasibility studies in diverse settings are advised to comprehensively show proof of concept, and randomised controlled trials methods may have value in terms of comparing the effect of PCVSA to usual care on key outcome measures, perhaps the timing of face-to-face GP visits, the identification of health problems, or the frequency of hospital admissions.

#### Methodological strengths & limitations

One of the key strengths of the study is the novelty of its aim to examine the feasibility and acceptance of PCVSA in general practice / primary care. Another strength of the study is its mixed-methods approach, combining both quantitative data and qualitative insights from semi-structured interviews with practice staff. This comprehensive evaluation provides a well-rounded understanding of the PCVSA’s impact on practice efficiency and patient engagement. However, there are limitations to consider. Firstly, being a pilot study, the study sample was small and specific to adult patients and staff in one practice. It is probable that the PCVSA would be less feasible and acceptable in other practice settings, particularly those with fewer support staff resources. Further testing of the study in larger and more diverse samples with varying service and patient characteristics is required to attain more generalisable findings. Also, although the quantitative survey captured patient satisfaction and perspectives, the study did not include a qualitative component from the patients’ perspective which could have provided deeper insights into their experiences and perceptions. Additionally, the study faced challenges with language barriers during the consenting process, which had a slight negative impact on recruitment and could have affected the clarity of communication and the overall participation experience for some patients with English language limitations that may have taken part. Further, GP visits were timed from the moment patients entered the GP’s office to the moment they left. While participants in this study did not raise this as an issue, future studies should note that GP times may be more accurately recorded if measurements also include time spent by the GP performing non-patient facing clinical duties.

## Conclusion

The study demonstrated that the PCVSA could be a valuable initiative in primary care and general practice. Based on the study’s findings, PCVSA may have significant potential for saving time, enhancing patient engagement, and facilitating more productive and efficient consultations. The findings suggest that further research in these settings is recommended to fully explore the benefits of integrating PCVSA into routine practice in primary care and general practice.

## Data Availability

Data availability statement This study’s quantitative raw data (i.e., PCVSA and GP consult timings, participant questionnaire responses) is accessible at https://zenodo.org/records/13952132. Data from a preliminary survey with 25 GPs in Ireland conducted by Wavescope Ltd. is also available at this website. The transcripts from this study’s qualitative interviews are not available online as publishing this data publicly is likely to breach the study participants’ confidentiality.

## Acknowledgements

We would like to thank the following for their support with this project: Enterprise Ireland, Centric Health, staff and patients at Ballyowen Medical (Dublin, Ireland), Afortiori Development, Xenon Health Solutions (Dublin, Ireland), Dr Louise Dubras (Ulster University, Belfast, Northern Ireland), Mr Seamus McGarrell (St James’s Hospital, Dublin Ireland), the Health Research Board (Dublin, Ireland), Ireland East Hospital Group, University College Dublin’s College of Health and Agricultural Sciences and School of Medicine.

## Supporting information captions

S1. Doctor Assessment Questionnaire

S2. Doctor satisfaction questionnaire

S3. Patient assessment questionnaire

S4. Qualitative interview topic guide

